# Breaking the back of COVID-19: Is Bangladesh doing enough testing?

**DOI:** 10.1101/2020.05.09.20096123

**Authors:** Hasinur Rahaman Khan, Tamanna Howlader

## Abstract

Following detection of the first 100 confirmed cases of COVID-19 in early April, Bangladesh stepped up its efforts to strengthen testing capacity in order to curb the spread of the disease across the country. This paper sheds light on the position of Bangladesh in relation to its South Asian neighbors India and Pakistan with respect to testing capacity and ability to detect cases with increased testing. It also analyzes recent data on case counts and testing numbers in Bangladesh, to provide an idea regarding the number of extra tests needed to detect a substantial number of cases within a short period of time. Findings indicate that compared to India and Pakistan, Bangladesh was able to detect more cases by increasing testing levels and expand its testing capacity by performing more per capita tests. In spite of these achievements, the rate of reported cases per 100 tests was consistently higher for Bangladesh compared to India, which suggests that in addition to increased testing, other factors, such as, effective enforcement of social distancing and efficient contact tracing are just as important in curbing the spread of the disease. The analysis reveals that current testing levels in Bangladesh are not adequate. Based on the findings, we recommend a 30-50% growth of the current test rate in the next few days so that by detecting and isolating more cases, Bangladesh could, in effect, contain the spread of new infections. The challenge, however, is to mobilize resources necessary to expand geographical coverage and improve testing quality while enforcing social distancing and performing efficient contact tracing.

## 1 Introduction

The severe acute respiratory syndrome coronavirus (SARS-CoV-2) pandemic fully established itself in Bangladesh in early April when the number of infected persons crossed the 100-th mark. Prior to this time, the country detected its first three coronavirus cases on 8 March 2020 confirmed by the Institute of Epidemiology, Disease Control and Research (Institute of Epidemiology and Disease Control and Research, 2020). The country detected three more cases of COVID-19 on March 16 taking the total number of infected persons to 8 (Khan & Hossain, 2020b). On March 18 the country witnessed its first COVID-19 death. Since then, the number of new infections continued to rise as Bangladesh ramped up the number of tests performed. By 5th of May, 93,405 tests had been performed as the disease spread to 63 of the 64 districts and the country counted 10229 cases and a death toll of 183 persons (Institute of Epidemiology and Disease Control and Research, 2020), (Khan, 2020).

Increasing the level of testing has been regarded as one of the most important tools in the fight against the disease. Testing leads to quick identification of cases and immediate isolation as well as treatment to prevent further spread. Once cases are identified, contact tracing may be performed to identify exposed individuals so that they too can be quickly isolated and treated if symptoms arise (Hellewell et. al., 2020). Thus, more and more testing helps to curb the spread and to flatten the curve and take the pressure off the health care system (Ravelo, 2020). In the case of South Korea, where there was high testing coverage, experts believe that testing played a major role in controlling the COVID-19 pandemic. The key to their success was a large, well-organized testing program combined with extensive efforts to isolate infected people and trace and quarantine their contacts. By 16th of March, South Korea had tested more than 270,000 people, many of whom were tested at a network of dozens of drive-through testing stations. This strategy, which has since been followed elsewhere, eased access to testing and prevented infected people from exposing others in waiting rooms (Cohen & Kupferschmidt, 2020). An additional advantage of widespread testing of “presumed COVID” patients who are not hospitalized is that it gives a far clearer picture of this new viral disease, which we currently have so little data about (Rosenthal, 2020). Thus, testing is important in the bigger public health picture on mitigation efforts, helping investigators characterize the prevalence, spread and contagiousness of the disease (Sanchez, 2020).

By May 5th, the USA administered the highest number of tests, i.e. over 7.5 million, which is over 12% of the global test total. Russia ranked second having conducted 4.6 million tests, Germany ranked third with over 2.7 million tests, and Italy ranked fourth having performed approximately 2.3 million tests up till May 5. Testing levels have differed widely across countries. Heterogeneity has been observed in testing capacity both within and between countries due to differences in factors such as financial resources, laboratory capacity and availability of qualified technicians. Healthcare systems in low and middle income countries such as Bangladesh have faced the greateat challenges in expanding their testing capacity due to limited resources.

Although there have been a number of research works on COVID-19 in Bangladesh, very few have focussed on testing. For instance, Khan & Hossain (2020a) analyzed global data and concluded that cumulative number of tests is not an important variable to predict the number of infections. Other studies on COVID-19 in Bangladesh include (Khan et al., 2020), (M. M. Islam et al., 2020), (Paul et al., 2020), (Mamun & Gri?th, 2020), (Shahidul et al., 2020), (Anwar et al., 2020), (M. D. Islam & Ayesha, 2020), (M. Islam et al., 2020). As of May 5, Bangladesh performed 567 tests per million people, while its South Asian neighbors India and Pakistan performed 864 and 1007 tests per million, respectively over the same period. A question that arises is, ’Has Bangladesh conducted an adequate number of tests to restrict the spread of COVID-19?’ In a country where majority of the people are poor and uneducated, enforcing social distancing measures can be challenging. Thus, adequate testing is required to identify and isolate cases to curb the spread of the disease, especially among the poor masses. This article attempts to answer the question of adequate testing by analyzing COVID-19 related data on Bangladesh uptil May 2, 2020 and evaluating the country’s position in relation to India and Pakistan.

## 2 Methodology

The data are collected from worldometers.info website (Max Roser & Ortiz-Ospina, 2020). We considered data reported as of May 2, 2020 for three countries Bangladesh, India and Pakistan. Furthermore the data pertaining to Bangladesh has been cross checked with the Bangladesh government’s source (Institute of Epidemiology and Disease Control and Research, 2020). Basic statistical tools for exploratory data analysis have been used.

## 3 Analysis

Figure 1 (top panel) shows a plot of the number of daily infections due to COVID-19 against number of days since the first 100 cases was identified in Bangladesh, India and Pakistan. The time series for Bangladesh is shorter than that for India and Pakistan because the first 100 cases were recorded earlier in the latter two countries. The three curves corresponding to the three countries demonstrate exponential increases in the number of daily infections. During the first 11 days, Bangladesh had nearly the same number of infections per day as Pakistan. Both countries exceeded the daily number of infections recorded in India. However after the 12th day, the number of daily infections recorded in Bangladesh began to rise above that of India and Pakistan. Although India had a lower number of daily case counts initially during the outbreak, the numbers began to climb after the 16th day overtaking Pakistan and Bangladesh after the 22nd and 26th days, respectively. The rapid increase in the slope of the curve for India relative to Pakistan during the later stages of the outbreak suggests that COVID-19 may be spreading more quickly in India. However, one must keep in mind that this graph does not take into account differences in the numbers of tests performed in the three countries.

**Figure 1:**
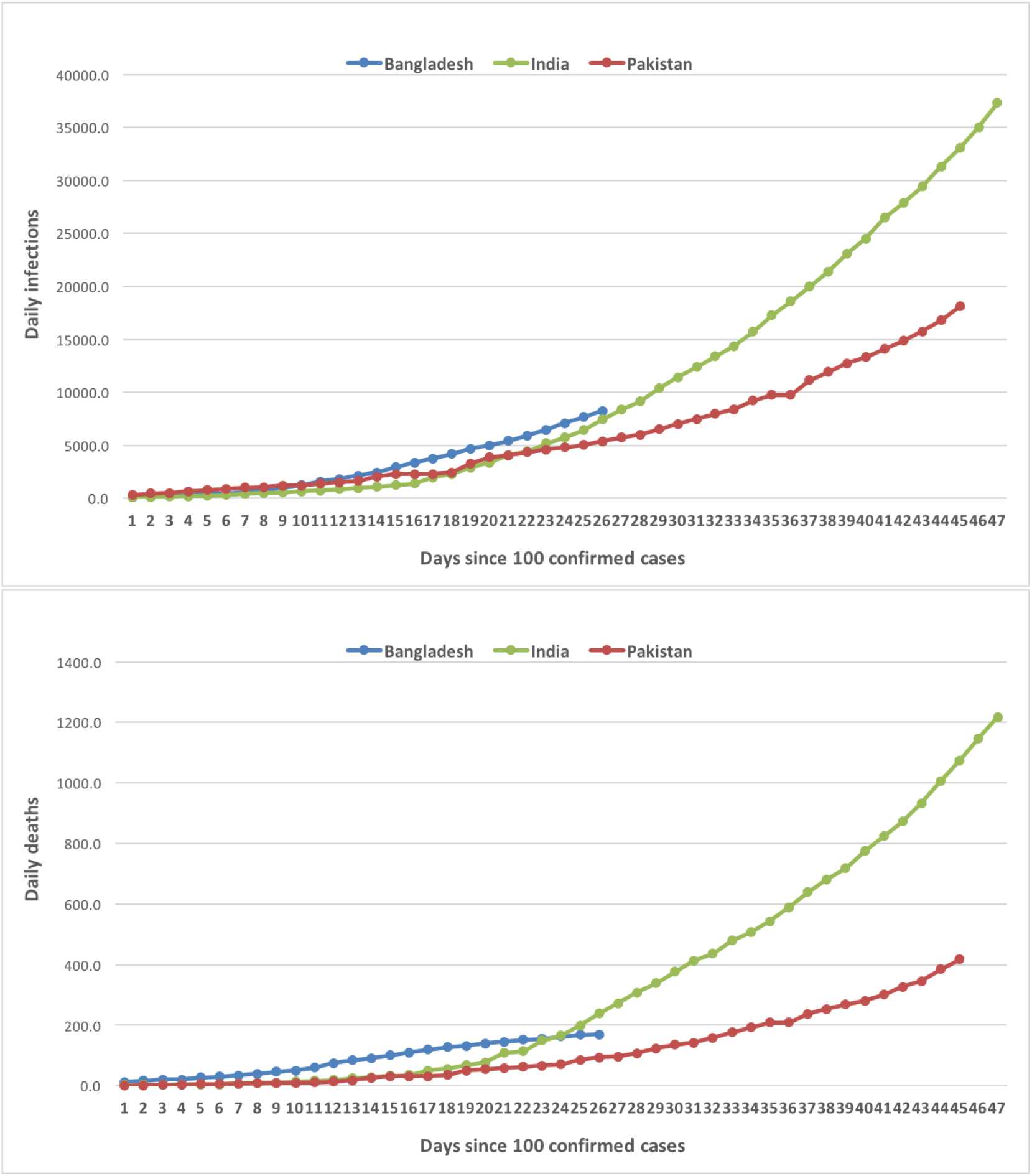
Daily infections and deaths of Bangladesh, India and Pakistan since their 100 confirmed cases

The plot in Figure 1 (bottom panel) shows the number of daily deaths against the number of days since 100 confirmed cases for Bangladesh, India and Pakistan. It can be seen that the number of recorded deaths was higher for Bangladesh compared to the other two countries from day 1. As the days progressed, the death toll in Bangladesh rose and was well above that of India and Pakistan after the 11th day. During the first 16 days, the curves for India and Pakistan overlapped remaining nearly flat for the first 11 days. However after the 16th day, the number of deaths in India picked up rapidly overtaking Bangladesh after the 23rd day. The rate of increase in daily deaths for India was higher than that of Bangladesh and Pakistan and continued to increase during this time.

Figure 4 indicates that there is a postive correlation between number of tests conducted and number of cases identified. Thus, the number of cases per 100 tests is a useful measure to compare disease frequency between countries because it controls for the number of tests that have been performed. Figure 2 shows the number of cases per 100 tests in the days following identification of the first 100 cases for Bangladesh, India and Pakistan. Compared to Bangladesh and Pakistan, the number of confirmed cases was lower for India, i.e. less than 10 cases per 100 tests, for most of the duration of the time series. Initially, Bangladesh also recorded less than 10 cases per 100 tests, but after the 10 th day, the number of cases began to rise and the ratio hovered between 10 to 20 cases per 100 tests. The time series for Pakistan was erratic during the first 24 days with several peaks exceeding 20 cases per 100 tests and one peak reaching as high as 70 cases per 100 tests. Over the next 14 days, the ratio remained within the 0-10 band and then jumped to slightly over 20 cases per 100 tests on the 39th day. It then declined and remained steady within the 10-20 band. Overall this graph, which gives an idea of the disease frequency after controlling for the number of tests performed, suggests that India may have been better able to control the spread of the disease compared to the other two countries.

**Figure 2:**
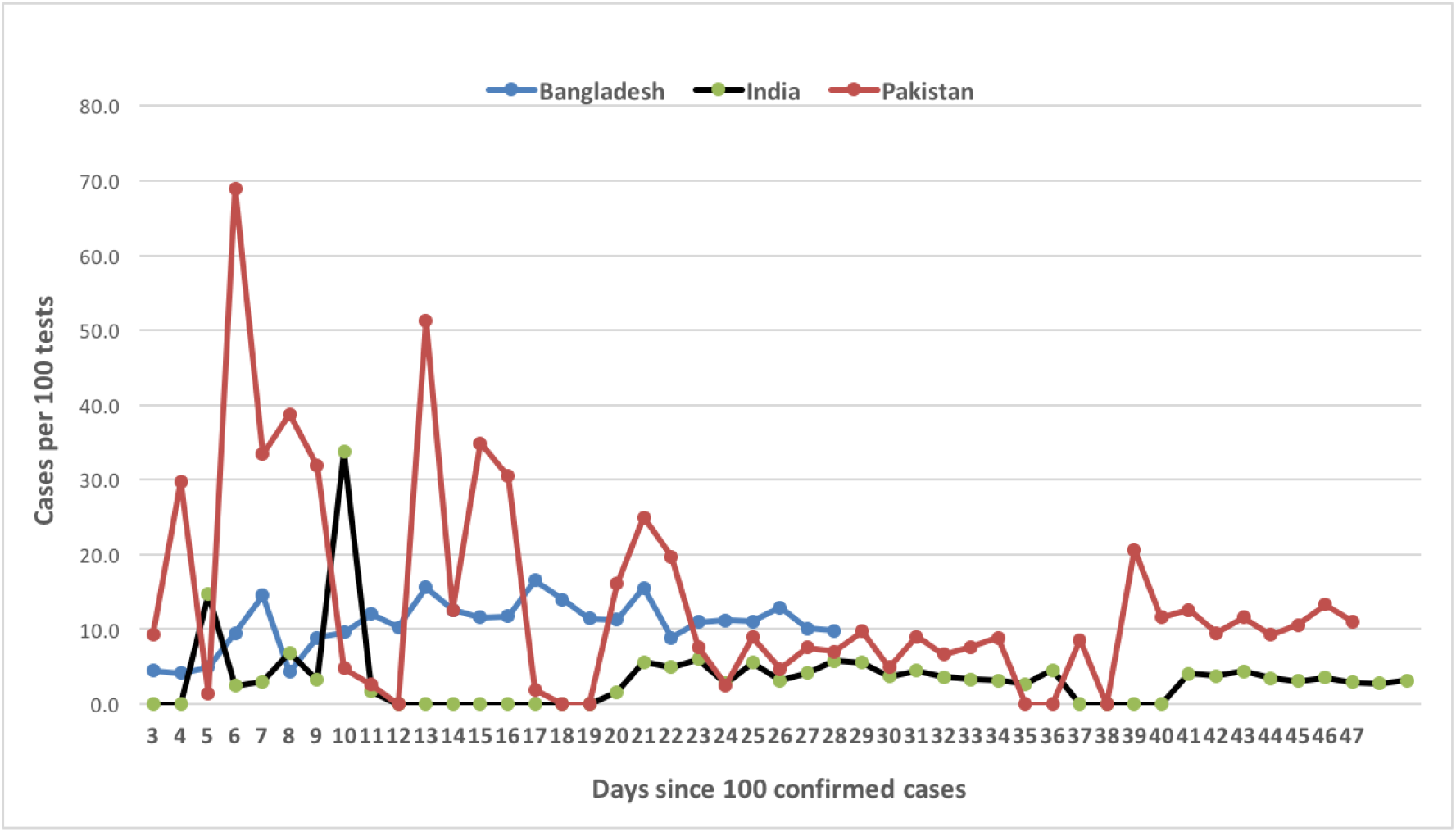
Confirmed cases per 100 tests of Bangladesh, India and Pakistan since their 100 confirmed cases

Testing is regarded as one of the most effective ways to deal with the rapidly speading COVID-19 disease because it gives authorities the opportunity to isolate infected cases and stem the spread of the disease. Figure 3 shows the number of tests performed per 1000 people against number of days since 100 confirmed cases for Bangladesh, India and Pakistan. There appears to be an exponential increase in the number of tests performed over time for all three countries. However, Bangladesh appears to have had the highest level of testing followed by Pakistan after controlling for population size. This seems to indicate that Bangladesh has done a good job at expanding its testing capacity compared to India and Pakistan.

**Figure 3:**
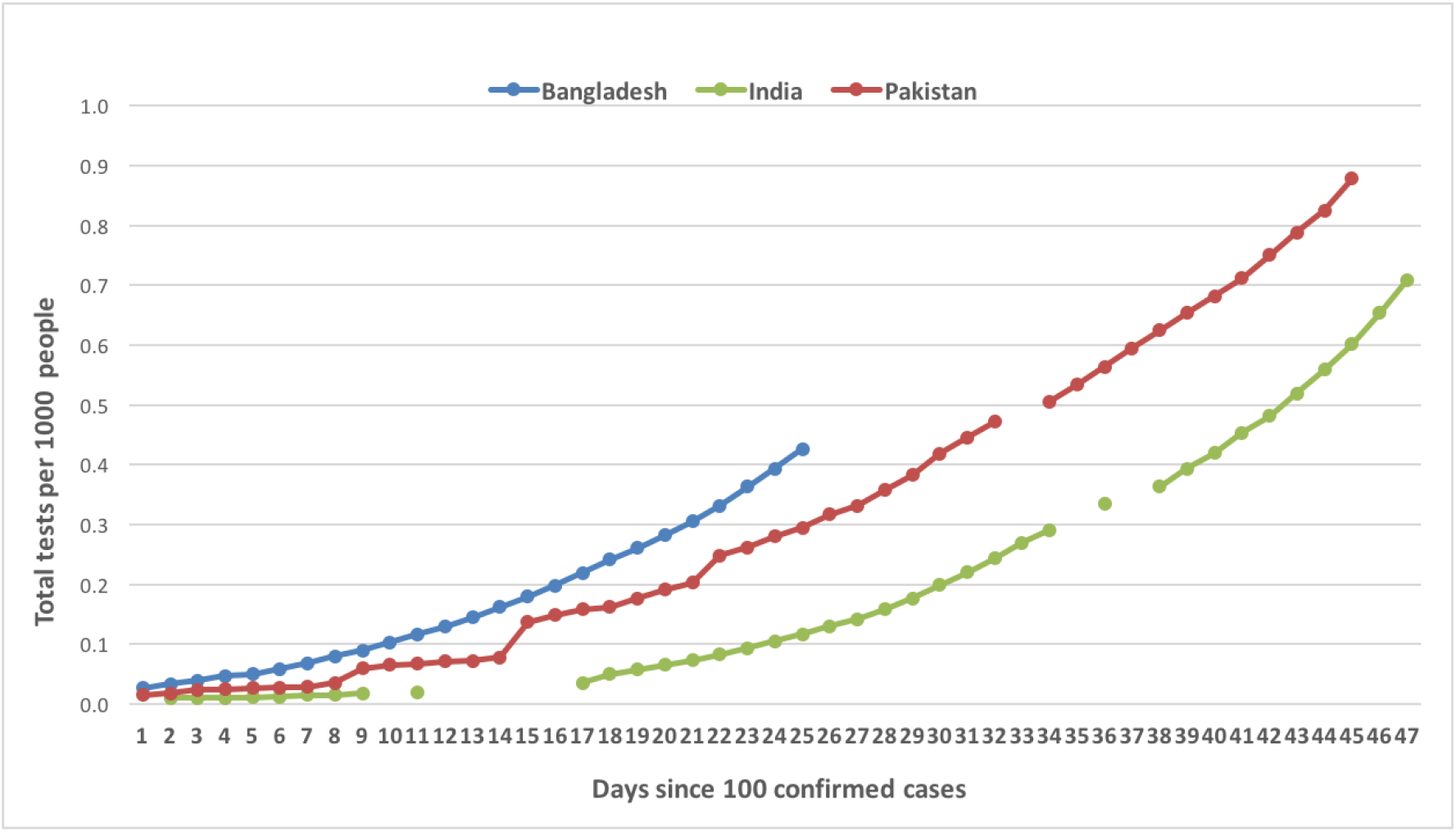
Number of tests per 1000 people of Bangladesh, India and Pakistan since their 100 confirmed cases

In terms of daily number of tests, Bangladesh downperforms India and Pakistan as found from the daily data (Max Roser & Ortiz-Ospina, 2020). But when the number of tests are adjusted by the country’s population, then Bangladesh outperforms India and Pakistan as shown in Figure (3). Is the current level of daily tests enough to slow down the spread? To search for an answer, we analysed the data in a way that provides some idea regarding the increment in the daily growth of tests needed to slow down the spread of the virus in Bangladesh. Table (1) shows the number of daily tests, number of cases, rate of confirmed cases in 100 tests, growth rate of the daily test in last 10 days, average of last 10 day’s daily rate of confirmed cases in 100 tests, and the minimum increment in the daily test rate that needs to be added to the next day’s test for slowing down the spread. The last four columns show the extra number of tests required and the resulting added number of infections, if testing were increased by 8.8%, which is the minimum requirement, and 20%. It appears from the analysis of last 14 days data from April 22 to May 5 that the case finding rate per 100 tests is nearly averaged at 12. That is, 12 cases are reported as positive in every 100 tests although a significant number of tests had been increased i.e. from 3772 (April 22) to 5573 (May 1)- which is 87.4% increment. In spite of such increments, the case finding rate was found to be fixed at around 12. This is true for other dates as well. Hence, this analysis suggests that Bangladesh had to increase number of tests by minimum 8.8% to lower the case finding rate. This finding will be persistent as long as a constant rate of confirmed case by the tests holds and the underlying sample collection strategies are uniformly maintained.

**Table 1:**
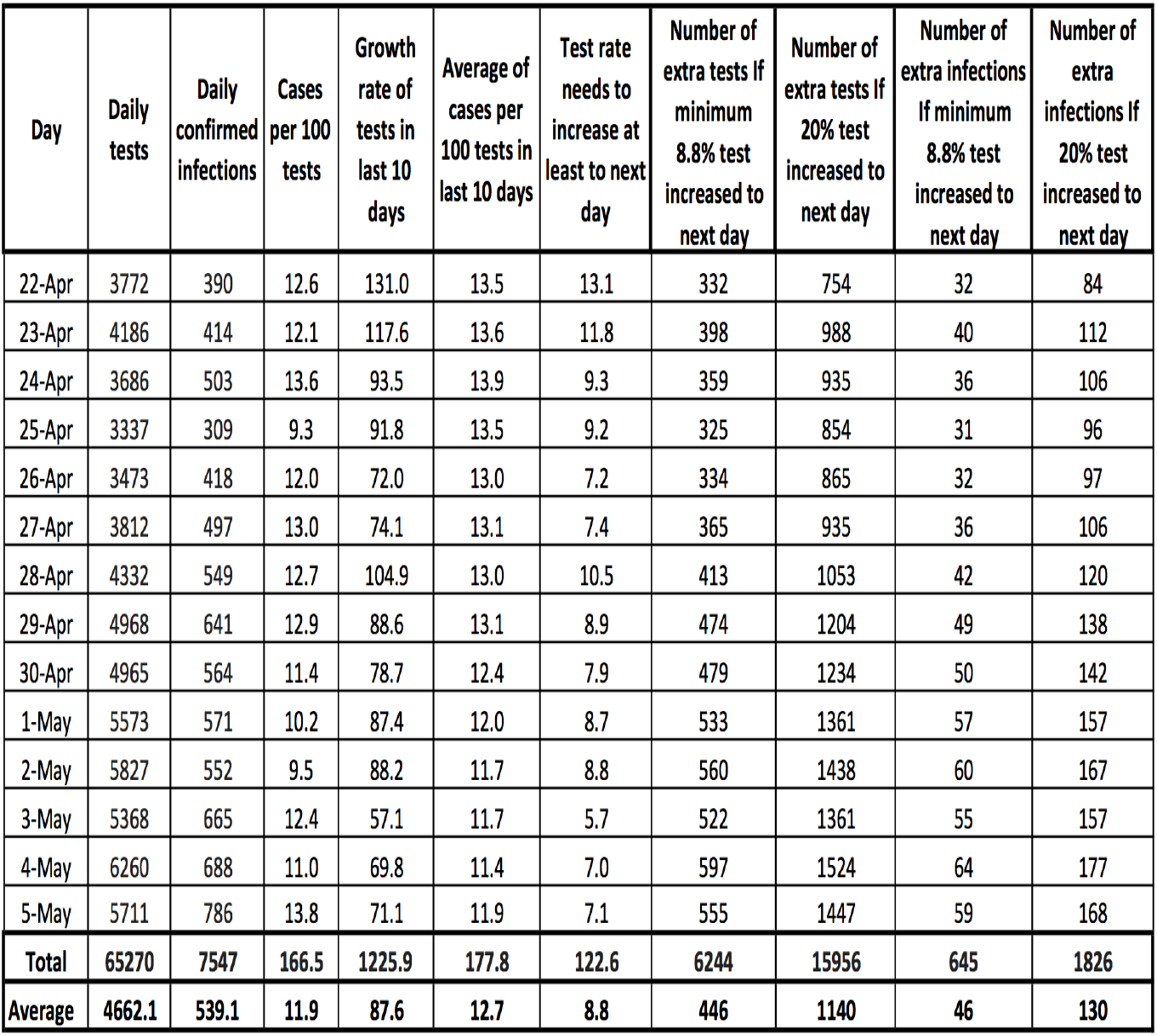
Dynamics of COVID-19 tests in Bangladesh

Table (1) shows also the predicted number of infections against the extra tests. It is found that if tests were increased every day at its minimum 8.8% rate then 6244 more tests could detect 645 infections during the period of April 22 to May 5. However, if this rate was increased by 20% then 15956 extra tests (with daily extra 1140) could be performed and in turn 1826 more infections (with daily extra 130) could be detected during that period.

It should be noted that there is very strong positive correlation (r=0.98) between the daily tests and the confirmed cases for Bangladesh while there is strong positive association (r=0.95) for India but there is just moderate positive association for Pakistan (r=0.64)(see Figure (4)). This also suggests that Bangladesh has consistently been able to detect higher number of cases with increased testing. Similar relationship between testing levels and case counts is observed for India although the country has observed slightly less consistency in detecting cases while Pakistan is far behind Bangladesh in this respect. Thus, although Bangladesh appears to be outperforming India and Pakistan in terms of greater case detection through increased testing, it needs to go for rapid testing expansion. A 30-50% growth or even doubling of the current test rate in next few days could help detect more cases and aid in controlling the virus spread within a short period of time.

**Figure 4:**
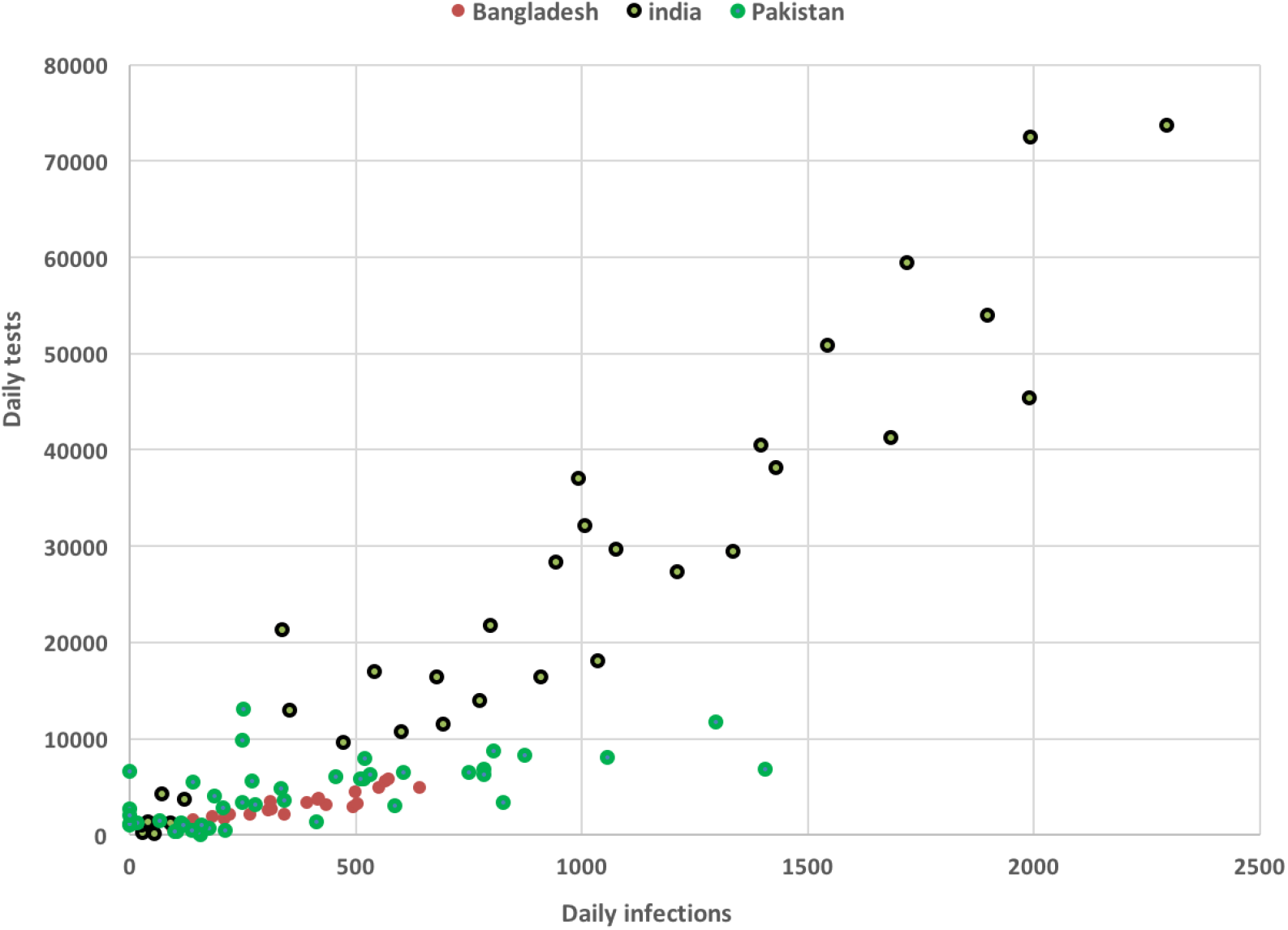
Scatter plot between the number of daily tests and confirmed cases for Bangladesh, India and Pakistan

## 4 Conclusions

Increased testing is seen as one of the most important strategies for combating the COVID-19 pandemic. The positive association between testing and the number of reported cases suggests that more cases will be identified and subsequently isolated with an increase in the number of tests performed. This is in turn will have the effect of slowing down the spread of the virus throughout the population. The objective of this paper has been to assess whether the current level of testing in Bangladesh is sufficient to curb the spread of the virus and to draw comparisons with two important South Asian countries, namely, India and Pakistan. Preliminary analysis revealed an exponential trend in daily case counts for all three countries during the first 26 days after the 100th case was found indicating that all three countries were in the initial stages of the epidemic.

Bangladesh had a higher daily death count than India and Pakistan till the 24th day since the first 100 cases were identified after which the death toll in India surpassed that of Bangladesh. Plots of number of cases per 100 tests against time revealed that Bangladesh had higher numbers of reported cases compared to India on average. However, Pakistan had higher values for this ratio on several occasions compared to Bangladesh including one occasion where the ratio was seven folds higher. The consistently lower levels of reported cases per 100 tests over time indicate that India may have been more successful in controlling the spread of the virus compared to Bangladesh and Pakistan. Graphs for total tests per 1000 people against time showed exponential increases in the level of testing for all three countries. However, the curve for Bangladesh was higher than that of India and Pakistan indicating that Bangladesh has been more successful in expanding its testing capacity with respect to its population size. In spite of this achievement, Bangladesh has observed higher numbers of reported cases per 100 tests than India, which seems to indicate that in addition to increased testing, other factors, such as, effective enforcement of social distancing and efficient contact tracing may also be important in curbing the spread of the disease.

This study also found that Bangladesh needed to increase every day a minimum 8.8% of tests to lower the case finding rate during April 22 to May 5. If tests were increased every day at its minimum 8.8% rate, then 6244 more tests could detect 645 infections during the same period. However, if this rate was increased by 20% then 15956 extra tests would need to be performed and this would lead to 1826 more infections being detected. We also found that Bangladesh has consistently been able to detect higher number of cases with increased testing. This has been observed for India as well although with slightly less consistency. For Pakistan, the correlation has not been as strong as for Bangladesh and India. In spite of outperforming India and Pakistan with respect to per capita testing levels, Bangladesh needs to go for rapid testing expansion, such as 30-50% growth or even doubling of the current figure in next few days to detect more cases. This, in effect, would help to curb the spread of the disease through greater contact tracing and isolation.

## Data Availability

The working data set used for this study has been submitted to the journal as additional supporting file.

https://ourworldindata.org/coronavirus

